# Perceptions towards mask use in school children during the SARS-CoV-2 pandemic: the Ciao Corona Study

**DOI:** 10.1101/2021.09.04.21262907

**Authors:** Priska Ammann, Agne Ulyte, Sarah R Haile, Milo A Puhan, Susi Kriemler, Thomas Radtke

**Author notes:** **Corresponding author** Thomas Radtke, PhD, Epidemiology, Biostatistics and Prevention Institute (EBPI), University of Zurich, Hirschengraben 84, 8001 Zürich, Switzerland. These authors have contributed equally to this work and share first authorship (Priska Ammann, Agne Ulyte). These authors have contributed equally to this work and share senior authorship (Susi Kriemler, Thomas Radtke).

## Abstract

**Background:** Mask wearing contributes to the reduction of transmission of severe acute respiratory syndrome coronavirus 2 (SARS-CoV-2). In Switzerland, compulsory use of masks was introduced in indoor public spaces and later in schools. In the canton of Zurich, masks were introduced for secondary school children (grades 7-9) from November 2020, and for primary school children (grades 4-6) from February 2021– along with other protective measures against SARS-CoV-2. This study explored perceptions towards the usefulness of masks in school and public in a cohort of children and adolescents in the canton of Zurich, Switzerland, in January – May 2021.

**Methods:** School children aged 10 to 17 years enrolled in *Ciao Corona*, a prospective school-based cohort study, responded to nested online surveys between January 12 to March 24 2021 (Q1) and March 10 to May 16 2021 (Q2). Secondary school children were surveyed at Q1 and Q2, and primary school children at Q2 only. Surveys for parents and their children included questions on children’s perception of the usefulness of masks and mask wearing behavior. Associations between perceived usefulness of masks and child’s school level, gender, and parents’ educational attainment were analyzed with Pearson’s and McNemar’s chi-squared tests. Free-text comments provided by children were classified into categories of expressed attitude towards mask wearing.

**Results:** 595 (54% girls) and 1118 (52% girls) school children responded to online questionnaires at Q1 and Q2, respectively. More than half of school children perceived masks to be useful at school (Q1:60% and Q2:57%) and in public (Q1:69% and Q2:60%). Girls perceived masks as useful more often than boys (at Q2 at school: 61% *versus* 53%, in public: 64% *versus* 57%), and children of parents with high educational attainment more often than those of parents with lower educational attainment (at Q2 at school: 61% *versus* 49%, in public: 63% *versus* 54%). There were no differences in the perceived usefulness of masks among children in primary versus secondary school. At Q1 and Q2 each, about 20% of children provided individual statements about masks, of which 36% at Q1 and 16% at Q2 reported side-effects and discomfort such as skin irritations, headache or difficulties breathing during physical education.

**Conclusion:** Approximately 60% of school children perceived masks at school and in public places as useful. A small but non-negligible proportion of children reported discomfort and side-effects that should be considered to ensure high adherence to mask wearing among school children.

**Trial registration:** ClinicalTrials.gov NCT04448717 https://clinicaltrials.gov/ct2/show/NCT04448717

**CONTRIBUTION TO THE FIELD STATEMENT:** Worldwide about 150 countries fully closed their schools at some point during the coronavirus pandemic, while other countries – such as Switzerland – kept schools open almost all the time. However, among other protective measures, children in secondary school (aged approximately 14-16 years) had to wear masks since November 2020, and older children in primary school (aged 11-13 years) – since February 2021.

As part of the large study *Ciao Corona* based in schools in Switzerland, we wanted to learn how children perceive the usefulness of masks in school and public. Children and their parents completed questionnaires in January-March (595 secondary school children) and March-May 2021 (1118 secondary and primary school children).

We found that about 60% of children perceived masks to be useful at school and in public. Girls perceived masks as useful more often than boys, and children of parents with university or college education more often than those of parents with lower education. About 7– 9% of children reported side-effects and discomfort such as skin irritations, headache or difficulties breathing during physical education. Although side-effects were not frequently reported, they should be considered to ensure high adherence to mask wearing among school children.

## INTRODUCTION

The contribution of school children to the spread of SARS-CoV-2 remains a controversial topic. In 2020-2021, policy decisions on schools ranged internationally from a complete closure over many months to keeping them fully open throughout the pandemic.^1,2^ While children rarely develop severe health outcomes ^3–5^, SARS-CoV-2 infection rates among children and adolescents are generally similar to those in adults.^6,7^ However, major outbreaks of clustered SARS-CoV-2 infections in schools are rare.^7–9^ Both the specific characteristics of SARS-CoV-2 infection in children (lower viral load and more frequent asymptomatic or mildly symptomatic infections) and the effective mitigation measures taken up by schools, including mask wearing of adults and children, could have contributed to the limited spread in schools.^10–12^ Despite this uncertainty, numerous countries implemented nationwide school closures to confine the diffusion of SARS-CoV-2 infections.^2,13^Worldwide, about 150 countries fully closed their schools at some point in 2020-2021, while only a few countries partially closed schools or kept schools open during the pandemic.^14^In Switzerland, schools were open with preventive measures in place, except during a 6-week nationwide lockdown from March 16 to May 10, 2020. Subsequently, schools were instructed to implement protective measures (e.g., social distancing, frequent hand washing, implementation of class bubbles in schools). Wearing masks became compulsory in public transport and in situations where physical distance (2m, later 1.5m) could not be maintained from July 2020 for all persons beyond 12 years age. From November 2020 on, masks became compulsory for adults and secondary school children (grade 7 and higher) in educational settings. From February 2021, wearing of masks was extended to primary school children in the middle school level (grade 4 and higher).

Although masks have been shown to reduce the risk of community SARS-CoV-2 transmission^15–19^, prolonged wearing of masks may be perceived as a burden and can be associated with potential side effects, such as headaches or skin irritation in adults and children.^20,21^ Willingness to wear masks is influenced by perceived benefits and social norms, and refusal by negative attitudes or side effects.^22,23^ To this date, the authors are unaware of any study investigating the perceived usefulness and attitudes towards mask wearing in children and adolescents. Hence, this study explored school children’s perceptions towards the usefulness of masks in- and outside the school setting from randomly selected schools in the canton of Zürich, Switzerland. In a second step, the study focused on the potential association between perception of usefulness and school level (secondary *versus* primary school), gender, and parents’ educational attainment^24^. In a third step, changes of perception over time were documented in secondary school children.

## METHODS

### *Ciao Corona* study

*Ciao Corona* is a prospective cohort study investigating SARS-CoV-2 seroprevalence in children and clustering of seropositive children within school classes in the Canton of Zurich, Switzerland. Details about the study design, main research questions and testing procedures can be found elsewhere.^7,25^ In brief, three testing phases (T1: June/July 2020, T2: October/November 2020, T3: March/April 2021) were completed between June 2020 and April 2021 including about 2500 school children aged 6 to 16 years longitudinally. At each testing phase, venous blood samples were taken for SARS-CoV-2 serologic analysis. Seroprevalence in children who were ever seropositive by T2 was 6.6% (95% credible interval (CrI) 4.0% to 8.9%), and increased to 16.4% (12.1% to 19.5%) by T3 (March/April 2021).^26^ Online questionnaires were distributed to parents to assess children’s symptoms, possible SARS-CoV-2 infections, and lifestyle. Parents of the participating children were asked to fill in a baseline questionnaire together with their child. They received further bi-monthly online follow-up questionnaires until March/May 2021.

Questions on mask wearing were asked at two time points: first, in the follow-up questionnaire in early 2021 (Q1: January 12 to March 24, 2021), and second, in late spring of 2021 (Q2: March 10 to May 16 2021; integrated into a follow-up for participants enrolled earlier, and the baseline questionnaire for participants enrolled in March-April 2021). Questions included children’s perception of the usefulness of masks in the school setting and in public, mask wearing practices (i.e., type of mask, storage, and wear time), and an open question for comments on masks (details in online supplementary material, Table S1). Questions on masks at Q1 were addressed only to secondary school children (grades 8-9, age 13-17 years), and to both primary (grades 5-6, age 10-13 years) and secondary school children at Q2 because masks were compulsory for primary school children of middle school level only from the end of January 2021.

### Data preparation and analysis

A total of 1302 of 2974 (44%) children who participated in at least one of the *Ciao Corona* testing rounds completed at least one question about masks in at least one of the Q1 and Q2 questionnaires. Variables extracted from the questionnaires were perceived usefulness of masks (i.e., 5-point Likert scale: 1: not at all useful, 2: somewhat useful 3: neither useful nor useless, 4: quite useful, and 5: very useful) at school and in public, school level (middle or upper), child’s gender and parents’ highest educational attainment as well as residence (urban/rural). For the analysis, the 5-point Likert scale used for the assessment of perceived mask usefulness was merged into three categories: “somewhat or not useful” (Likert scale values 1-2), “neutral” (Likert scale value 3), and “useful” (Likert scale values 4-5). High educational attainment was defined as at least one parent having at least a technical college or university degree.

Two authors (PA, TR) developed six categories for the free-text comments by identifying homogenous themes in the free text responses: 1) general rejection and frustration; 2) general acceptance; 3) side effects and discomfort; 4) uncertainty and mistrust; 5) situation-specific complaints (e.g., during sports); and 6) not categorizable. These two authors independently categorized free-text responses to the open question on masks into the defined categories. Discrepancies in categorization were discussed among the two authors and consensus achieved through discussion.

Descriptive statistics included summary of participants’ key characteristics (age, gender, school level, rural or urban residence, and parents’ highest educational attainment). Associations between school level, gender, parents’ highest educational attainment and perceived usefulness of masks were investigated with Pearson’s and McNemar’s chi-squared tests. In addition, we examined the differences in perceived usefulness between the two measurement points Q1 and Q2 on a population (cross-sectional) and individual (longitudinal) levels. Data from Q2 was used for the main cross-sectional analyses, with data from Q1 presented in the Supplemental material. Data analysis was performed with R version 4.1.0 (R Core Team, 2020).^27^

## RESULTS

### Study population characteristics

Participant characteristics are given in Table 1. 595 (54% girls) study participants responded to at least one question on masks in the online survey at Q1, and 1118 (52% girls) participants at Q2. At Q2, 596 middle level school children and 522 upper level school children completed the questionnaire. The majority of children had at least one parent with a Swiss nationality, and at least one parent with a higher educational attainment (Table 1).

**Table 1.** Participant characteristics

|  | January – March 2021<br>(Q1) | April – May 2021<br>(Q2) |
| --- | --- | --- |
|  | *N* = 595 | *N* = 1118 |
| Age (years) | 14.3±0.7 | 12.8±1.6 |
| Gender, n (% girls)* | 320 (54%) | 580 (52%) |
| School location (urban), n (%) | 239 (46%) | 427 (46%) |
| RT-PCR confirmed SARS-CoV-2 infection, n (%)** | 21 (4%) | 43 (4%) |
| **School level \*\*\*** |  |  |
| Middle level (grades 5–6) | - | 596 (53%) |
| Upper level (grades 8–9) | 595 (100%) | 522 (47%) |
| **Socioeconomic status** |  |  |
| Either parent with Swiss nationality, n (%) | 503 (85%) | 974 (87%) |
| Either parent with high educational attainment, n (%) | 349 (60%) | 783 (71%) |

Data are mean±standard deviation (SD) or count (%). RT-PCR, real time polymerase chain reaction; SARS-CoV-2, severe acute respiratory syndrome coronavirus 2.
\* 3 (Q1) and 2 (Q2) children reported “other” as their gender.
\*\*RT-PCR confirmed SARS-CoV-2 infection was defined as at least one RT-PCR positive test reported in any questionnaire by 24 March 2021 (Q1), or by 16 May (Q2).
\*\*\*For school children, mask use was compulsory from October 29, 2020, for grade 7 and higher, and from January 25, 2021, for grade 4 and higher.

### Perception of mask usefulness

At Q1, the majority of children and adolescents perceived masks as useful, both at school (n = 357, 60%) and in public (n = 408, 69%). At Q2, 57% (638) participants perceived masks as useful at school, and 60% (675) in public. At Q2, masks were perceived as useful at school by 354 (59%) of primary and 284 (55%) of secondary school children (p=0.2), and in public by 366 (61%) of primary and 309 (59%) of secondary school children (p=0.2). Detailed responses to the questions of mask usefulness at Q1 and Q2 for both school levels are provided in Supplementary Table S2.

At Q2, girls perceived mask wearing as useful more often than boys both at school (61% vs 53%, p = .018; Figure 1A) and in public (64% vs 57%, *p* = .026); Figure S1). The difference between girls and boys was similar at Q1 (at school: 63% vs 56%, p=.2; in public: 71% vs 67%, p=.3; Supplementary Table S3).

**Figure 1.**
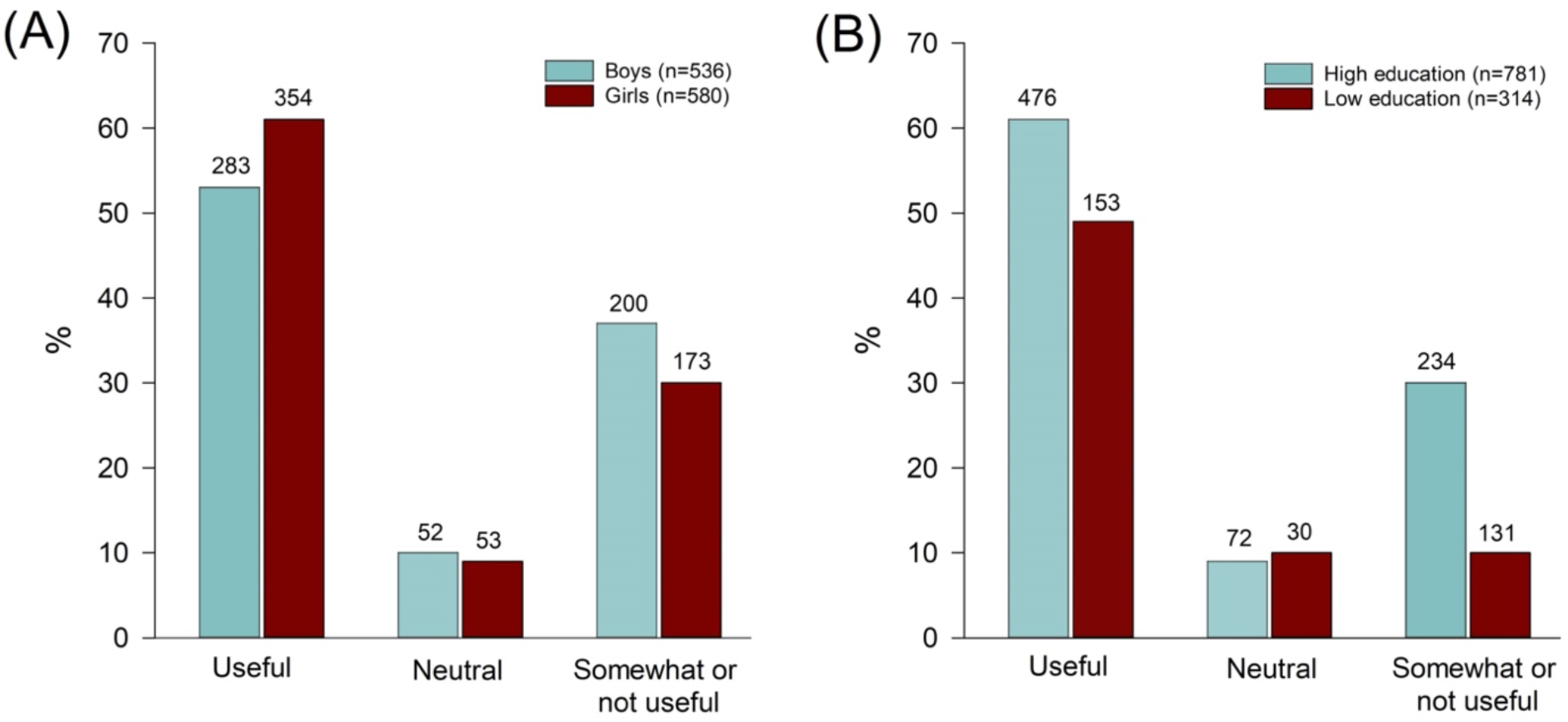
Comparison of perceived usefulness of mask wearing at school between boys and girls (A) and according to parents’ educational attainment (B) at Q2 Two children indicated their gender as “other”. Higher educational attainment was defined as at least one parent having at least a technical college or university degree.

At Q2, 71% (781 of 1095 with reported information) of participating children had at least one parent with high educational attainment. Children of parents with higher educational attainment were more likely to perceive mask wearing as useful at school (61% vs 49%, *p* < 0.001; Figure 1B) and in public (63% vs 54%, *p* =0.019; Figure S1). At Q1, the difference was somewhat smaller for perceived usefulness at school (63% vs 57%, p=0.3), and similar to that in Q2 for usefulness in public (74% vs 63%, p=0.026; Supplementary Table S3).

### Longitudinal comparison among secondary school children between Q1 and Q2

From the secondary school children participating at Q1, 75% (446 of 595) answered the question on perceived usefulness of mask wearing at school and 75% (447 of 595) on mask wearing in public at Q2. Overall, the proportion of children perceiving masks as useful was smaller at Q2 than at Q1 (*p*_school_ = 0.06, *p*_public_ < 0.001; Table 2). 17% of children reported a lower perception of mask usefulness at school at Q2 than at Q1, and 12% reported a higher perception of mask usefulness. 23% reported a lower perception of mask usefulness in public at Q2 than at Q1, and 10% reported a higher perception of mask usefulness.

**Table 2.** Comparison of perceived usefulness of mask wearing at school (n = 446) and in public (n = 447) between secondary school children at Q1 and Q2

| School | Q2 |  |  |  |
| --- | --- | --- | --- | --- |
| Q1 | Useful | Neutral | Somewhat or not useful | *p*-value* |
| Useful | 206 (46%) | 20 (4%) | 46 (10%) | 0.06 |
| Neutral | 13 (3%) | 5 (1%) | 11 (2%) |  |
| Somewhat or not useful | 26 (6%) | 14 (3%) | 105 (24%) |  |
| Public |  |  |  |  |
| Useful | 230 (52%) | 21 (5%) | 66 (15%) | < 0.001 |
| Neutral | 10 (2%) | 5 (1%) | 14 (3%) |  |
| Somewhat or not useful | 23 (5%) | 11 (2%) | 67 (15%) |  |

\* McNemar's chi-squared test.

### Free-text comments on masks

A total of 120 (20%) children provided a free-text comment about masks at Q1, and 228 (20%) at Q2. Children who perceived mask wearing as somewhat or not useful both at school and in public were more likely to leave a comment, both at Q1 (*p*_public_ = 0.02, *p*_school_ < 0.001), and Q2 (*p*_public_ = 0.006, *p*_school_ = 0.001). At Q1, 95 (80%) comments were categorized as either general or situation-specific mask rejection, complaints or mistrust, in comparison to 140 (61%) such comments at Q2. Most comments mentioned side effects and discomfort [36% at Q1 (7% of all children), 18% at Q2 (4% of all children)] and general rejection/frustration (23% at Q1, 21% at Q2). Table 3 shows some examples of comments about masks for each of the six defined categories.

**Table 3.** Examples of free-text comments on an open question on mask wearing at Q1 and Q2.

| Response category and comment examples* | Q1<br>*N* = 120 | Q2<br>*N* = 228 |
| --- | --- | --- |
| **General rejection and frustration** | 28 (23%) | 48 (21%) |
| • “Masks are annoying”<br>• “Hopefully, there will soon be a life without masks!”<br>• “I hate masks”<br>• “All the students are annoyed by the masks” |  |  |
| **Uncertainty and mistrust** | 12 (10%) | 10 (4%) |
| • “Masks make no sense, everything is just fear mongering”<br>• “Completely useless”<br>• “Why do we have to wear masks in class but not in school sports?<br>There, the distance can also not be maintained...”<br>• “Masks are useless against viruses and have no effect on<br>infection!” |  |  |
| **General acceptance** | 15 (13%) | 37 (16%) |
| • “For me, wearing masks in everyday life makes a lot of sense.<br>The only thing that bothers me are people who think it is not<br>important, because they believe they won’t get sick. In school, I<br>find that wearing masks would need to be better controlled,<br>because many do not follow the rules or see each other at lunch<br>without a mask and share food or drinks.”<br>• “They do not look nice, but are currently the least evil, so there is<br>worse.” |  |  |
| **Side-effects and discomfort** | 43 (36%) | 41 (18%) |
| • “I get too little air, so that I have headaches more often.”<br>• “My skin is very unclean. I miss seeing the whole face of people<br>around me.”<br>• “Because of the masks, I have skin rashes and my ears hurt.”<br>• “I think that wearing masks is very bad. You can hardly breathe,<br>and I can concentrate less and less in school. I find this idea very<br>bad, because in your free time children talk to each other without<br>a mask anyway”. |  |  |
| **Situation-specific complaints** | 12 (10%) | 41 (18%) |
| • “Very annoying while doing sports!!!!”<br>• “It’s unbearable in gym class!”<br>• “It is very uncomfortable to breathe during sports” |  |  |
| **Not categorizable** | 10 (8%) | 51 (22%) |
| • “More fabric masks”<br>• “No” |  |  |

\*Individual responses were translated from German.

### Mask type, storage and daily wear time

The majority of children reported using disposable masks (69% at Q1 and 76% at Q2) or a combination of disposable and fabric masks (21% at Q1 and 15% at Q2) (see Supplementary Table S4). When not worn, masks were most commonly carried in a pocket or fastened around the arm, and less frequently stored in a plastic or fabric bag. Mean reported daily wear times of masks were about 6 hours for middle level school children, and 7 hours for upper level school children (Table S4).

## DISCUSSION

This study explored the perception of the usefulness of masks at school and in public among children and adolescents from randomly selected schools enrolled in the *Ciao Corona* study, from January to May 2021 in the canton of Zurich, Switzerland. We found that approximately 60% of participants perceive wearing of masks as useful, both at school and in public. Girls valued the usefulness of masks higher than boys, and children of at least on parent with a higher education (i.e., technical college or university degree) perceived masks as more useful compared to those of parents of lower educational background. Perceived usefulness of wearing masks in public, but not at school, decreased over time. Among all survey participants, a small proportion of school children reported general side effects and discomfort (7% at Q1 and 4% at Q2) or situation-specific complaints (2% at Q1 and 4% at Q2) associated with masks.

In this study, we found a 60% acceptance of wearing masks with a large proportion of school children reporting to perceive them as useful at school (57-60%) and in public (60-69%). Some survey participants providing free-text comments about masks reported on the necessity of wearing masks to protect others, and some children even complained about others not wearing their masks according to school rules. On the other hand, roughly one third of the survey participants did not perceive masks in school-settings as useful. Among those, participants provided specific free-text comments about masks significantly more frequently, indicating a bias towards negative comments. Such comments were sometimes rather non-informative (“masks are useless”, “I hate masks”), but others pointed towards physical discomfort and side-effects, such as fatigue, decreased concentration, headaches, or acne. Mandatory wearing of masks was often criticized for specific situations, especially during physical education classes but also during breaks.

Complaints about masks, particularly of their side-effects, need to be considered on an individual basis. In settings with potential widespread transmission and where physical distance of at least one meter cannot be guaranteed, the use of masks for children aged 6 years and older is recommended by the World Health Organization (WHO) and the United Nations Children’s Fund (UNICEF).^28^ As schools were fully open in Switzerland during the SARS-CoV-2 pandemic, except for a 6-week nationwide lockdown, the wearing of masks was introduced for school children grades 4 and higher along with other protective measures to keep transmission rates as low as possible. Importantly, the current evidence for the use of masks during the SARS-CoV-2 pandemic in the pediatric population including benefit–harm evaluation is limited. Laboratory-based experimental studies investigating short-term effects of various types of masks (e.g., surgical mask, FFP2, or N95 respirators) worn at rest and during exercise on physiological variables and individuals’ perception of breathing are controversial.^29–31^ Long-term studies and benefit-harm assessments of the burden of masks and other mitigation measures, complete closures of schools ^32–34^, and effect on SARS-CoV-2 transmission mitigation would be needed to evaluate the role of masks in the pediatric population. Yet, as masks are usually only one of several implemented mitigation measures, it will always be challenging to tease out the unique effect of masks on infection control.

Children whose parents had a higher educational background and girls perceived masks as more useful. In contrast, we did not see a difference between perception of middle and upper school level children, as a proxy of age. Associations between gender and education with the willingness to wear masks has been shown in previous studies in adults.^24,35,36^ For example, in an analysis of almost 10’000 shoppers, female gender, older age, and urban location were associated with higher odds of an individual to wear a mask.^24^ Hence, schools could provide tailored information about masks to reach different groups of children, and to actively engage them in a dialogue about potential benefits and harms.

Although the absolute difference was small, the proportion of school children rating masks as useful in public was higher than the proportion rating masks useful at school (60% *versus* 57% at Q2). One potential explanation could stem from the prolonged wearing of masks in school-settings where masks, on average, are worn between six and eight hours a day, as reported by participants (Supplementary Table S4). Outside the school-setting, masks are usually only required short-term in certain places, such as public transport or supermarkets. Secondly, children and adolescents may feel safer in a classroom environment than in public due to the implementation of various additional measures as part of a school’s protection concept, and thus perceive mask wearing there as relatively less useful. In addition, the proportion of children perceiving mask wearing as useful slightly decreased from January to April 2021, at school from 60% to 57% and in public from 69% to 60%, respectively. This could be related to the longer overall exposure of wearing masks at Q2 compared to Q1, as well as to the lower community incidence of SARS-CoV-2 infections at Q2.

This study has a number of limitations. First, the online questionnaires nested within the *Ciao Corona* study were usually completed by parents/legal guardians. Parents were explicitly encouraged to complete the part of the questionnaires about masks together with their children or to let their child respond to these questions alone. Nevertheless, it is not entirely clear to what extent children’s perceptions towards masks were reported by children themselves, and thus captured correctly, as social desirability bias is possible. Second, children’s behaviour and perceptions are largely influenced by their parents’, teachers’, and peers’ beliefs and behaviour.^37,38^ Thus, it is also likely that parents who were skeptical towards certain restrictions and measures implemented during the SARS-CoV-2 pandemic did not take part in the *Ciao Corona* study, and did not allow their children to participate, resulting in potential selection bias in the study population. Third, side-effects reported by some survey participants cannot be (clinically) verified and would require a different study design. Finally, our study sample consisted of a larger proportion of parents with high educational background (60% at Q1 and 71% at Q2) than would be expected in the general population.^39^ Consequently, our findings likely do not reflect the entire spectrum of perceptions about the usefulness of masks in the general population, and may to some extent overestimate the perceived value of masks in schools and public.

## CONCLUSION

In summary, our findings suggest that 60% of children perceive masks as a useful measure to mitigate SARS-CoV-2 transmission at school and in public, with slightly higher perception of usefulness among girls and families of higher socio-economic background. Certain complaints of some children, such as physical discomfort and side-effects due to the prolonged wearing of masks, should be addressed in order to ensure the maintenance of mask wearing, and ultimately protect the health and well-being of children and adults at schools and beyond.

## Data Availability

The raw data supporting the conclusions of this article will be made available by the authors, on reasonable request.

## AUTHOR CONTRIBUTIONS

SK and MAP initiated the cohort study in which this study is embedded. AU, PA, TR and SK designed the research questions and the study, and were responsible for the study organization and procedures. PA was responsible for the organization of testing at schools in 2021. PA and TR were responsible for the online surveys and provided guidance to participants, if needed. AU, SRH, and PA carried out the statistical analysis. All authors contributed to the interpretation of study findings. PA and TR wrote the first draft of the manuscript, and all authors critically reviewed and approved the final version of the manuscript.

## ACKNOWLEDGMENTS

We kindly thank all children and their parents for their participation in this study and for completing the online surveys.

## ETHICS APPROVAL

The study was approved by the Ethics Committee of the Canton of Zurich, Switzerland (2020-01336). All participants provided written informed consent before being enrolled in the study.

