## Supplementary files for "Perceptions towards mask use in school children during the SARS-CoV-2 pandemic: the Ciao Corona Study"

**Supplemental Table S1** Questions related to mask wearing at Q1 (follow-up questionnaire) and Q2 (follow-up questionnaire and baseline questionnaire for new participants)

| Mask-related questions | Response categories |
|---|---|
| How useful do you think it is to wear masks in public spaces (outside of school)? | 1 – not at all useful, 2 – somewhat useful 3 – neither useful nor useless, 4 – quite useful, 5 – very useful |
| How useful do you think it is to wear masks at school? | 1 – not at all useful, 2 – somewhat useful, 3 – neither useful nor useless, 4 – quite useful, 5 – very useful |
| How many hours a day do you wear a mask? | ______ hours per day. |
| How often do you change the mask? | 1 – after each wearing (>1 hour), 2 – after four hours of wearing, 3 – 1-2x daily, 4 – daily, 5 – every few days to weekly, 6 – never/only when I lost it |
| How do you store your mask? | 1 – loosely in the pant pocket/handbag/tied around the arm, 2 – in a plastic bag, 3 – in a paper bag, 4 – in a cloth bag |
| What type of mask are you wearing? | 1 – disposable mask (purchased), 2 – cloth mask (bought), 3 – cloth mask (homemade) |
| Anything else you want to tell us about masks? | |

Note: questions and response categories were translated from German.

**Supplemental Table S2** Perceived mask usefulness at Q1 and Q2 ratings

| | Q1 | Q2 | | |
|---|---|---|---|---|
| | ML (N = 595) | Total (N = 1118)* | ML (N = 596) | UL (N = 522)** |
| **Mask usefulness at school** | | | | |
| 1 – not useful at all | 91 (15%) | 157 (14%) | 78 (13%) | 79 (15%) |
| 2 – a little useful | 107 (18%) | 217 (19%) | 108 (18%) | 109 (21%) |
| 3 – neither useful nor useless | 40 (7%) | 105 (9%) | 56 (9%) | 49 (10%) |
| 4 – rather useful | 216 (36%) | 365 (33%) | 218 (37%) | 147 (28%) |
| 5 – very useful | 141 (24%) | 273 (25%) | 136 (23%) | 137 (26%) |
| **Mask usefulness in public** | | | | |
| 1 – not useful at all | 49 (8%) | 106 (9%) | 54 (9%) | 52 (10%) |
| 2 – a little useful | 91 (15%) | 228 (20%) | 112 (19%) | 116 (22%) |
| 3 – neither useful nor useless | 47 (8%) | 108 (10%) | 64 (11%) | 44 (8%) |
| 4 – rather useful | 243 (41%) | 379 (34%) | 215 (36%) | 164 (32%) |
| 5 – very useful | 165 (28%) | 296 (27%) | 151 (25%) | 145 (28%) |

ML: middle school level (grades 5–6) in primary school, UL: upper school level (grades 8–9) in secondary school.

* From them, 1117 children answered the question on mask usefulness at school, and 1117 on mask usefulness in public.

** From them, 521 children answered the question on mask usefulness at school, and 521 on mask usefulness in public.

**Supplemental Table S3** Comparison of perceived usefulness of mask wearing at school and in public between boys and girls, and according to parents' educational attainment at Q1

| | Gender | | | Educational attainment of parents | | |
|---|---|---|---|---|---|---|
| **Usefulness at school** | **Male** **$N$ = 272** | **Female** **$N$ = 320** | $p$-value* | **Lower** **$N$ = 230** | **Higher** **$N$ = 349** | $p$-value* |
| useful | 152 (56%) | 203 (63%) | | 131 (57%) | 220 (63%) | |
| neutral | 19 (7%) | 21 (7%) | 0.2 | 15 (7%) | 24 (7%) | 0.3 |
| Somewhat or not useful | 101 (37%) | 96 (30%) | | 84 (37%) | 105 (30%) | |
| **Usefulness in public** | | | | | | |
| useful | 180 (66%) | 227 (71%) | | 146 (63%) | 257 (74%) | |
| neutral | 21 (8%) | 26 (8%) | 0.3 | 23 (10%) | 21 (6%) | 0.026 |
| Somewhat or not useful | 71 (26%) | 67 (21%) | | 61 (27%) | 71 (20%) | |

**Supplemental Table S4** Mask type, storage and wear time

| | Q1 | Q2 | |
|---|---|---|---|
| | **UL (N = 595)** | **ML (N = 596)** | **UL (N = 522)** |
| **Type of mask** | | | |
| Disposable mask | 411 | 421 | 429 |
| Fabric mask – purchased | 54 | 62 | 36 |
| Fabric mask – self-made | 3 | 1 | 2 |
| Combination of several mask types | 127 | 112 | 55 |
| **Storage of mask** | | | |
| Pocket/around the arm | 348 | 317 | 346 |
| Plastic bag | 108 | 150 | 84 |
| Paper bag | 41 | 36 | 21 |
| Fabric bag | 30 | 25 | 25 |
| Combination of several storages | 68 | 68 | 46 |
| **Daily wear time** (hours) | 7.4 ± 1.4 | 6.3±1.9 | 7.4±1.9 |
| | | ML and UL: 6.8±1.9 | |

ML: middle school level (grade 4-6), UL: upper school level (grade 7-9). Data are numbers (%).

**Supplemental Figure S1** Comparison of perceived usefulness of mask wearing in public between boys and girls (A) and according to parents' educational attainment (B) at Q2.

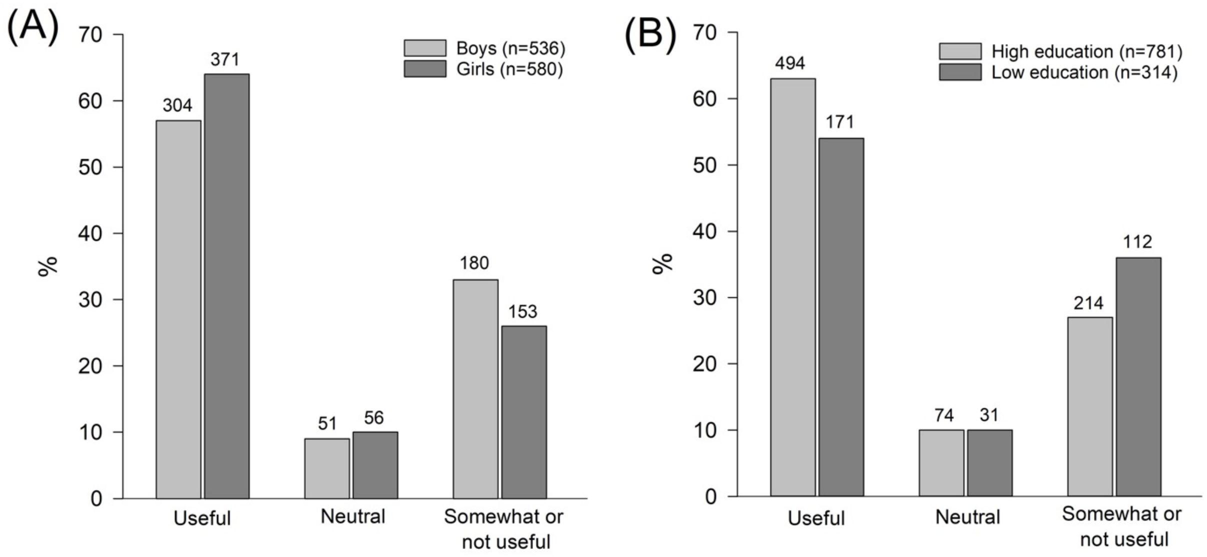


The distribution of responses was statistically significantly different between boys and girls (p = .026, Figure S1 A), and children with parents with higher and lower educational attainment (p = .019, Figure S1 B).
